# Prevalence of *Plasmodium vivax* in a semi-arid region of northern Kenya

**DOI:** 10.1101/2023.02.28.23286362

**Authors:** Wendy Prudhomme O’Meara, Linda Maraga, Hannah Meredith, Daniel Esimit, Gilchrist Lokoel, Tabitha Chepkwony, Joseph Kipkoech, George Ambani, Diana Menya, Elizabeth Freedman, Steve Taylor, Andrew Obala

## Abstract

Most malaria morbidity in Kenya is due to *Plasmodium falciparum* with no cases attributed to *P.vivax*. Little is known about the epidemiology in northern Kenya along the border with Ethiopia and Sudan. We found that 2% of household members of *P.falciparum* cases were infected with *P.vivax*, affecting all ages in urban and rural sites.

## Introduction

Until recently, little to no endemic transmission of *Plasmodium vivax* was reported in sub-Saharan Africa (SSA) outside of the Horn of Africa(1). *P*.*vivax* was presumed to be largely absent in the region owing to the scarcity of the Duffy blood group antigen. Over the last five years, however, accumulating evidence of endemic *P*.*vivax* has led to the recognition that it may be present in many areas of SSA, albeit at low levels, and Duffy-negative individuals can be infected and contribute to onward transmission(2, 3).

Turkana County is the northwestern most county in Kenya and shares a border with Uganda, South Sudan, and Ethiopia. The harsh climate is characterized by an average rainfall less than 215 mm per year and daytime temperatures of 40C. Malaria transmission in the region was predicted to occur in isolated pockets with epidemic potential only after unusual rainfall. However, reactive case detection conducted by our team across central Turkana documented year-round symptomatic and asymptomatic *P. falciparum* infections and confirmed perennial endemic transmission of malaria(4). Given stable malaria transmission and proximity to Ethiopia where *P. vivax* is endemic, we hypothesized that *P. vivax* may also be circulating.

## Methods

To test this, we extracted genomic DNA from 3305 dried bloodspots collected from household members of *P. falciparum* cases enrolled from three rural and three urban health facilities in Central Turkana(4). We tested each for the presence of *P*.*vivax* using an established nested PCR protocol(5).Gel electrophoresis bands were called by two observers independently.Fifteen extracts were randomly selected to be re-tested using the same primer sequences in a probe-based real-time PCR assay(6) and all were confirmed. For this analysis, we use the results from the first round of testing based on the nested qualitative PCR assay.

## Results

Two percent (2.1%, 69/3305) of household members were infected with *P. vivax*. Of those, nearly half (45%, 31/69) were co-infections with *P. falciparum* (**Table**). *P. vivax* was detected across our study transect throughout most of the year (**Figure)** with the highest prevalence recorded near an urban facility in Lodwar town (5.8%, 28/485). Infections were present across all ages with a slightly higher proportion of *P*.*vivax* mono-infections in children under 5 years (1.6%,Table). Ten *P*.*vivax*-infected participants reported malaria-like symptoms when they were screened and seven of those were coinfected with *P*.*falciparum*. Only three *P*.*vivax* infected participants had a malaria-like illness in the last month and none of these reported taking antimalarials. None of the *P*.*vivax*-infected participants reported travel in the preceding two months outside of their subcounty and 11 (16%) reported having a net for their sleeping space, slightly less than the proportion of uninfected participants having a net (19.7%).

## Discussion

From a clinical perspective, the burden of *P. vivax* in SSA remains unclear. *P. vivax* infections in SSA are rarely diagnosed in a clinical setting and may often be asymptomatic. The recommended rapid diagnostic test (RDT) for most SSA countries is *P. falciparum*-specific. Consequently, the burden of *P. vivax* may be underestimated or undocumented.

From the perspective of malaria elimination efforts, the implications of endemic *P. vivax* transmission are manifold. Strategies designed to target *P. falciparum* are undermined by *P. vivax* owing to the difficulty of detecting and curing dormant hypnozoites which can relapse and sustain transmission(7). *P. vivax* infections generate gametocytes before symptoms present, making it challenging to detect and treat before onward transmission occurs. Thus, *P. vivax* could present a growing challenge, even as *P. falciparum* is brought under control, as has been observed in co-endemic settings in Southeast Asia(8).

*Anopheles stephensi* was recently collected for the first time in Kenya(9), and the potential expansion of this highly competent vector of both *P. vivax* and *P. falciparum* into SSA could dramatically enhance the transmission of *P. vivax. An. stephensi* has been newly detected in five countries in SSA in the last 10 years. It is estimated that continued spread of this invasive vector would put an *additional* 126 million people at risk for malaria(10). The identification of *An. stephensi* in Djibouti was linked with a >100-fold rise in malaria cases, including the <u>first autochthonous cases of</u> *<u>P. vivax</u>* in 2016(11). If this emerging, outdoor-biting species becomes established across Kenya in the presence of confirmed *P. vivax* cases, it could dramatically increase the difficulty of malaria elimination in Kenya. Enhanced surveillance for both *An. stephensi* and *P. vivax* in areas at risk will inform the application of control measures as well as the allocation of clinical resources to enable safe treatment of cases of vivax malaria.

**Figure 1.**
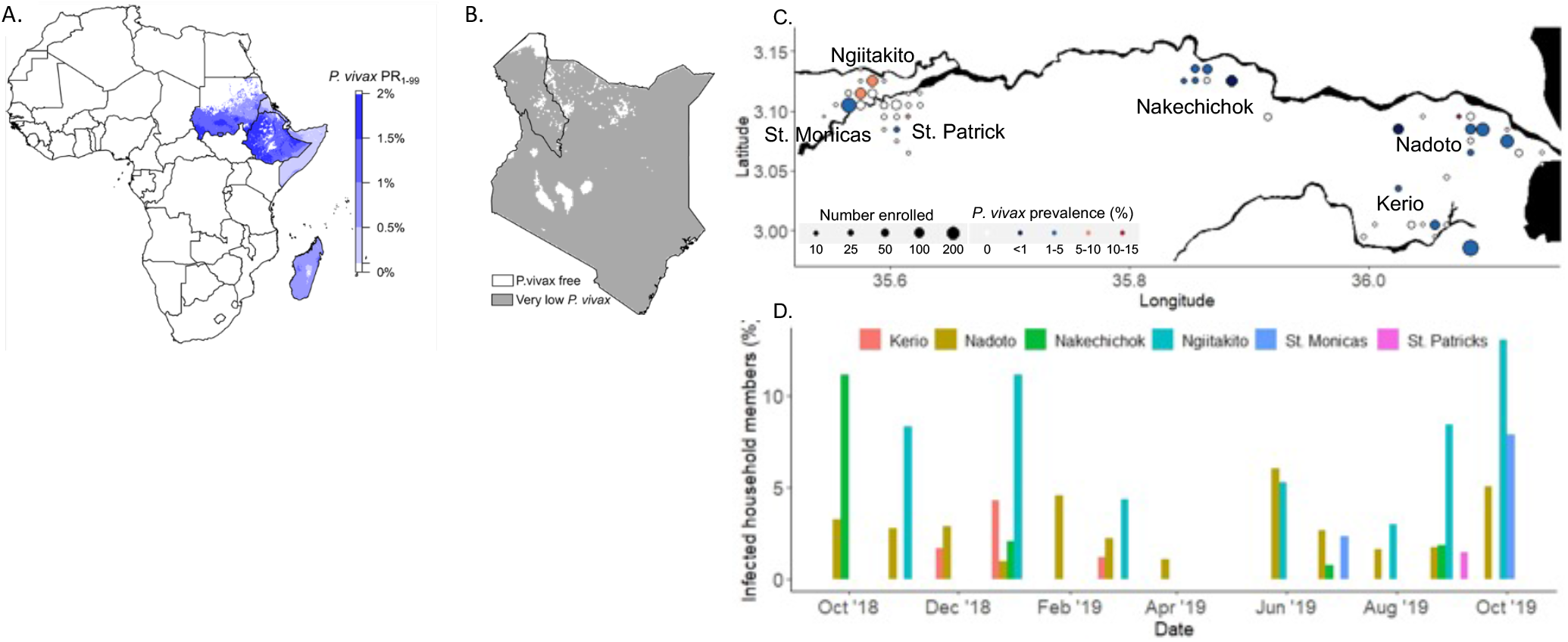
Estimates of *P. vivax* endemicity (2019) across Africa (**A**) and Kenya (**B**). Grey color is less than 0.01% prevalence, indicating transmission is not excluded by climate, but is very rare or has not been detected. By contrast, neighboring Ethiopia has established *P*.*vivax* transmission with a population prevalence of up to 2%. Turkana County border is outlined. Data from the Malaria Atlas Project. (C) Prevalence of P. vivax infection in communities along the Turkwel river where households of P.falciparum cases were tested for P.vivax infection. (D) P.vivax cases by month and health facility catchment

**Table 1:** *P. falciparum* and *P. vivax* infection by age. Results from 3201 tested for both *P. falciparum* and *P*.*vivax* where the participant age was recorded are shown. Forty-seven tested samples were missing age information, of which 20 were infected with P. falciparum and none with *P*.*vivax*. Fifty-seven samples were tested for *P*.*vivax* but not *P*.*falciparum*, of which none were positive for *P*.*vivax. P*.*falciparum* PCR methods and results are reported in (4).

|  | Age in years |  |  |  |
| --- | --- | --- | --- | --- |
|  | <5 | 6-15 | 16-40 | >40 |
| Any Plasmodium | 33.9% (166/490) | 34.4% (368/1069) | 30.0% (397/1324) | 30.8% (98/318) |
| *P. falciparum* only | 30.8% (151/490) | 32.2% (344/1069) | 28.2% (373/1324) | 28.9% (92/318) |
| *P. vivax* only | 1.6% (8/490) | 1.2% (13/1069) | 0.98% (13/1324) | 1.3% (4/318) |
| Mixed | 1.4% (7/490) | 1.0% (11/1069) | 0.83% (11/1324) | 0.63% (2/318) |

## Data Availability

All data produced in the present study are available upon reasonable request to the authors

